# INTerrupting prolifERation of Carbapenem resistance in Indonesia: clinical and genomic Evaluation of Pathways of Transmission (INTERCEPT): a Study Protocol

**DOI:** 10.64898/2026.08.28.26361608

**Authors:** Helmia Farida, Rebriarina Hapsari, Endang Sri Lestari, Nur Farhanah, Adam P. Roberts, Fabrice E. Graf, Russel Dacombe, Maria Moore, Joseph M. Lewis

**Affiliations:** Department of Pediatrics, Faculty of Medicine, Universitas Diponegoro, Semarang, Indonesia; Department of Microbiology, Faculty of Medicine, Universitas Diponegoro, Semarang, Indonesia; Department of Internal Medicine, Faculty of Medicine, Universitas Diponegoro, Semarang, Indonesia; Department of Tropical Disease Biology, Liverpool School of Tropical Medicine, Liverpool, United Kingdom; Department of Clinical Sciences, Liverpool School of Tropical Medicine, Liverpool, United Kingdom; Department of International Public Health, Liverpool School of Tropical Medicine, Liverpool, United Kingdom

**Keywords:** Carbapenem-resistant organisms, genomic epidemiology, plasmid transmission, transmission pathways, hospital transmission, community transmission, infection prevention and control, low-and middle-income countries

## Abstract

**Background:** Carbapenem-resistant bacteria are a major global public health threat, classified as critical priority pathogens by the WHO. In Indonesia, despite a national antimicrobial resistance control programme established by the Ministry of Health in 2015, resistance rates continue to rise, including increasing carbapenem resistance among clinically important bacteria. Strengthening approaches to directly interrupt transmission is essential, yet transmission pathways remain poorly understood with limited research and policy guidance within the Indonesian context.

**Methods and analysis:** The INTERCEPT study is a UK-Indonesia multidisciplinary collaboration aiming to identify transmission routes of carbapenem-resistant bacteria across healthcare and community settings, and the mechanisms of resistance gene transfer between bacteria and mobile genetic elements. We will conduct genomic surveillance of hospital inpatients, healthcare workers, hospital environments, and surrounding communities, including wastewater systems, combined with genomic analyses and mathematical transmission modelling. A cohort of patients with bloodstream infections will be recruited to evaluate resistant bacteria, treatment practices, and clinical outcomes. Qualitative research will explore behavioural and system-level factors influencing transmission and intervention implementation. Findings will inform stakeholder workshops to co-design context-specific interventions, with pilot intervention over 9 months with pre-and post-intervention assessment to guide scalable strategies to reduce AMR transmission.

**Discussion:** The INTERCEPT study addresses carbapenem resistance in Indonesia using an integrated approach combining microbiological surveillance, genomics, modelling, and qualitative methods. Strengths include cross-sectoral analysis (patients, workers, environment) and participatory intervention design. Limitations include geographic scope restricted to Central Java, Indonesia.

**Trial/study registration:** The INTERCEPT study has been prospectively registered in the ISRCTN registry, a WHO-recognised primary clinical study registry (ISRCTN36830327)

## Introduction

Antimicrobial resistance (AMR), associated with an estimated five million deaths annually worldwide, is a major global public health threat (1). Among resistant pathogens, bacteria producing carbapenem-inactivating enzymes are classified by the World Health Organization as critical priority pathogens due to their limited treatment options and high associated mortality (2). The burden of carbapenem resistance is particularly high in many low-and middle-income countries (LMICs), where access to effective alternative therapies is limited and healthcare systems face significant constraints.

In Indonesia, a middle-income country with a large and diverse healthcare system, the situation is increasingly concerning. National and global surveillance data indicate that carbapenem resistance is common among key pathogens, with reported resistance rates of 66% in *Acinetobacter baumannii* and 24% in *Klebsiella pneumoniae* bloodstream isolates (3). Despite the establishment of a national antimicrobial resistance control programme by the Ministry of Health in 2015, resistance rates continue to rise. While antimicrobial stewardship and infection prevention and control (IPC) programmes have been implemented, their effectiveness in reducing transmission remains uncertain (4), and data describing the epidemiology, clinical impact, and transmission dynamics of carbapenem-resistant organisms in Indonesia remain limited.

Existing evidence suggests that nosocomial transmission plays a major role in the spread of carbapenem-resistant bacteria. For example, studies from tertiary-care intensive care units in Jakarta indicate that a substantial proportion of patients acquire colonisation during hospital admission. However, data from other healthcare settings, including secondary and lower-level hospitals and the surrounding community, are scarce. Furthermore, colonisation—widely considered a precursor to invasive infection (5)—remains poorly characterised in this context, as do the factors driving transmission between patients, healthcare workers, and the environment.

The emergence and spread of carbapenem resistance in Indonesia reflect earlier patterns observed with extended-spectrum beta-lactamase (ESBL)-producing organisms. Widespread use of third-generation cephalosporins, particularly ceftriaxone, has likely driven high prevalence of ESBL-producing bacteria, which in turn has increased reliance on carbapenems (6). This cycle of escalating antimicrobial use and resistance has occurred despite the implementation of stewardship and IPC interventions, suggesting that current approaches may be insufficient to interrupt transmission at scale. There are knowledge gaps surrounding bacterial transmission within the Indonesian healthcare system and a critical need to develop locally adapted strategies that target transmission pathways.

Transmission of antimicrobial resistance is a complex, multi-level process occurring across interconnected levels, including genes, bacteria, patients, healthcare workers, the hospital environment, and the wider community (Figure 1). At the microbiological level, resistance genes can be transferred within and between bacterial species via mobile genetic elements (Figure 1C-D); these bacteria and resistance genes can be transmitted between people (Figure 1B) and between healthcare facilities and the community as people navigate the healthcare system (Figure 1A). Understanding these processes requires integrated approaches that combine clinical, microbiological, genomic, and behavioural data. The INTERCEPT study integrates data across these levels to identify key transmission pathways and aims to address these knowledge gaps through a comprehensive, multidisciplinary investigation of carbapenem resistance transmission in Indonesia.

**Figure 1.**
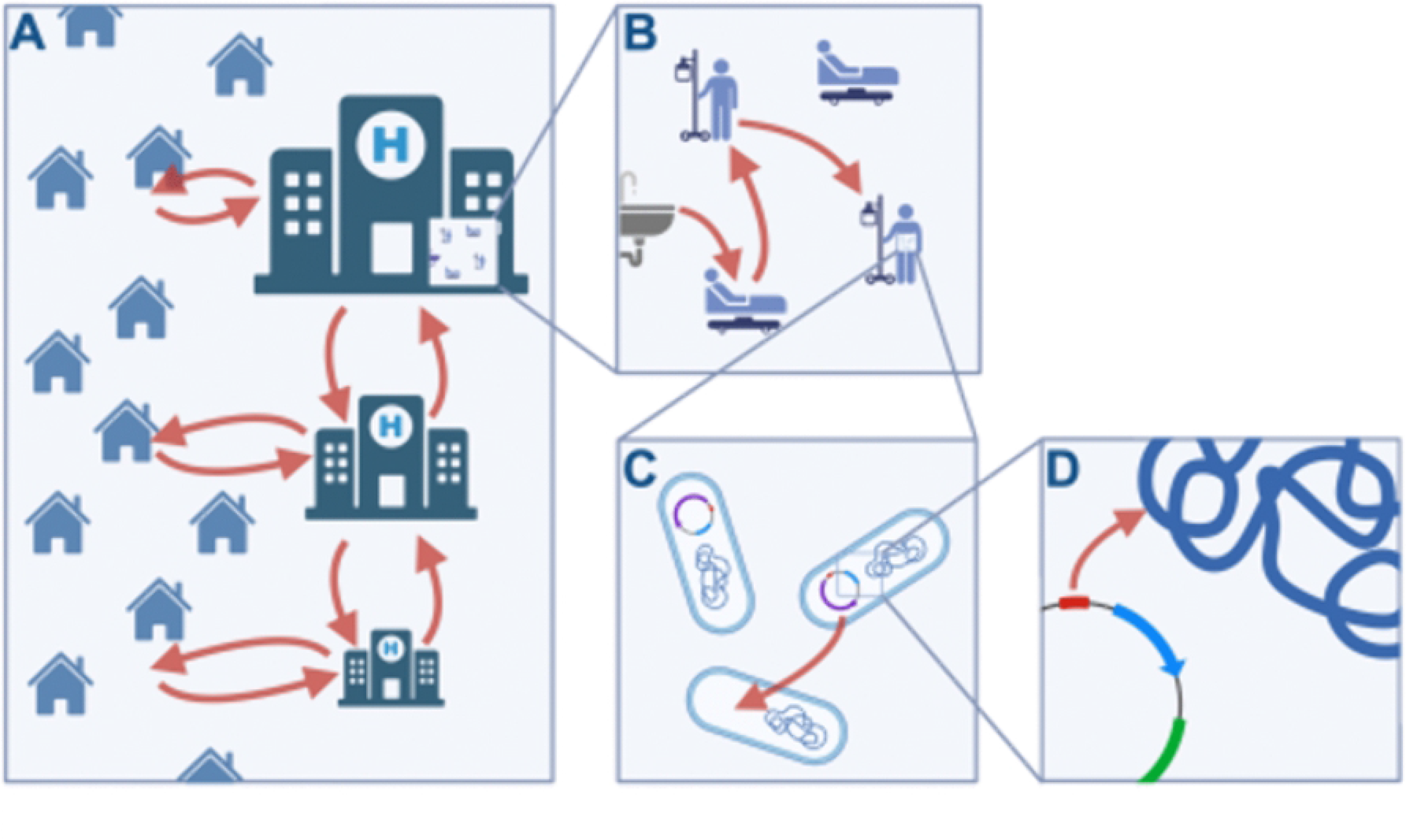
Conceptual framework of carbapenem resistance transmission across healthcare and community settings.

Specifically, the study will: (1) characterise transmission pathways across healthcare and community settings through systematic sampling of humans and the environment; (2) define the burden, treatment, and outcomes of carbapenem-resistant bloodstream infections; and (3) identify behavioural and system-level barriers to effective implementation of IPC and antimicrobial stewardship interventions. Using whole-genome sequencing, mathematical transmission modelling, and laboratory-based analyses, the study will provide high-resolution insights into the movement of carbapenem-resistant organisms and carbapenem resistance genes within and between different levels of the healthcare system.

Findings from this work will inform the co-design and pilot implementation of locally appropriate interventions aimed at interrupting transmission. In addition to generating evidence to support national AMR control strategies, INTERCEPT will strengthen local research capacity in genomics, bioinformatics, and qualitative methods, supporting sustainable responses to emerging resistance threats in Indonesia.

## Methods

### Study design

The INTERCEPT study is a multi-component, multidisciplinary research programme comprising three linked sub-studies designed to investigate transmission, clinical impact, and prevention of carbapenem-resistant bacteria in Indonesia.

First, INTERCEPT-Transmission, an observational cohort study of hospital inpatients and community members, will integrate systematic sampling of inpatients, healthcare workers, hospital environments, and hospital wastewater to characterise colonisation dynamics and identify transmission pathways. Concurrent antimicrobial point prevalence surveys will be conducted to describe antimicrobial use at study sites.

Second, INTERCEPT-Bacteraemia is an observational cohort study of patients with Gram-negative bloodstream infections, which will use anonymised routinely collected clinical data to determine the clinical impact, treatment patterns, and outcomes associated with carbapenem-resistant bloodstream infection.

Third, INTERCEPT-Intervention, a qualitative and interventional study, will explore stakeholder perspectives on antimicrobial stewardship and infection prevention and control (IPC), identify behavioural and system-level barriers, and support the co-design and pilot implementation of context-specific interventions to interrupt transmission.

These sub-studies will be conducted across three hospitals. Data collection will occur in rotating 6-week sampling periods at each site, repeated across wet and dry seasons to account for seasonality. The full sampling cycle will be conducted before and after implementation of a 9-month pilot intervention, enabling comparison of pre-and post-intervention conditions to assess impact.

### Study settings

The study will be conducted across three hospitals representing different levels of the Indonesian healthcare system: a tertiary referral hospital, a secondary referral hospital, and a district-level hospital. These facilities are located in urban and semi-urban settings in Central Java, Indonesia, and serve populations with diverse demographic and healthcare access characteristics.

All participating hospitals are accredited under the Indonesian national hospital accreditation system and have established IPC programmes and AMR control teams, although the level of implementation and available resources vary between sites. The hospitals differ in size, bed capacity, and critical care facilities, reflecting the heterogeneity of healthcare delivery across the Indonesian system.

The inclusion of hospitals across multiple levels of care enables the study to capture patient flow, antimicrobial use, and transmission dynamics across the healthcare network, as well as interactions between hospitals and the surrounding communities.

### Study procedures

#### Recruitment procedures, eligibility, and consent

The INTERCEPT study will use a rolling recruitment strategy across the three participating hospitals. Sampling will be conducted sequentially at each site during a dedicated 6-week sampling period and repeated in both the wet and dry seasons. The full sampling cycle (i.e. two 6-week sampling periods at each of the three participating hospitals – a total of 36 weeks or around 9 months) will be performed before and after implementation of pilot interventions to be able to assess the impact of the interventions.

Eligibility criteria were defined separately for each of the three INTERCEPT sub-studies.

For the INTERCEPT-Transmission study, participants will comprise three groups with inclusion and exclusion criteria as detailed in Table 1. Community participants with recent hospitalisation (within the previous six months) will be excluded to minimise misclassification of healthcare-associated colonisation (Table 1).

**Table 1:** Inclusion and exclusion criteria for INTERCEPT-substudies.

| Sub-study | Inclusion criteria | Exclusion criteria |
| --- | --- | --- |
| INTERCEPT-Transmission | Arm 1: | Arm 1 |
|  | Adults (18+) admitted to participating wards | Undergoing management with palliative intent and/or likely to die within 24 hours<br>Does not speak English or Indonesian<br>Lack capacity to consent to enrolment<br>AND no representative can be identified |
|  | Arm 2: | Arm 2: |
|  | Community-dwelling adults (18+) in the community served by the | Hospital admission within previous 6 months (to minimise misclassification of |
|  | hospital | healthcare-associated colonisation). |
|  |  | • Lacks capacity to consent to enrolment<br>• Does not speak English or Indonesian |
|  | Arm 3 | Arm 3 |
|  | Staff members (adults 18+) working on participating wards | • Not willing or available to participate in the study<br>• Does not speak English or Indonesian |
| INTERCEPT-Bacteraemia | All patients (adults and children) with Gram-negative bacilli identified in blood cultures | None |
| INTERCEPT-Intervention | Adult stakeholders (≥18 years) involved in infection prevention and control (IPC) and/or antimicrobial stewardship activities within participating hospitals or at local and national policy levels. | • Not willing or available to participate in the study<br>• Does not speak English or Indonesian |

For the INTERCEPT-Bacteraemia study, all patients (adults and children) with Gram-negative bacilli identified in blood cultures will be eligible (Table 1). No additional exclusion criteria will be applied.

For the INTERCEPT-Intervention study, eligible participants will include adult stakeholders (≥18 years) involved in IPC and/or antimicrobial stewardship activities within participating hospitals or at local and national policy levels. Individuals who are unwilling or unable to participate, or unable to communicate in Indonesian or English, will be excluded (Table 1).

For the INTERCEPT-Transmission study, three participant groups will be recruited: hospital inpatients, community members, and healthcare workers. Hospital inpatients admitted to participating wards during the recruitment period will be screened for eligibility and approached by the study team. At each hospital, there will be two participating wards: one ICU and one general ward. Community participants will be recruited from areas surrounding participating hospitals using household-based sampling approaches. Healthcare workers from participating wards will also be invited to participate.

For the INTERCEPT-Bacteraemia study, eligible participants will be identified through routine microbiology laboratory service at participating hospitals. Patients with Gram-negative bacilli identified in blood cultures during the recruitment period will be included using anonymised routinely collected clinical data.

For the INTERCEPT-Intervention study, purposive and snowball sampling strategies will be used to recruit stakeholders involved in antimicrobial stewardship and IPC, including clinicians, nurses, pharmacists, hospital managers, policymakers, and support staff. Focus group discussions will involve frontline ward staff, while ethnographic observations will be conducted in participating wards.

Written informed consent will be obtained from all participants prior to enrolment, except in the INTERCEPT-Bacteraemia study, which will use anonymised routinely collected clinical data without direct participant contact. Participants will receive written and verbal explanations regarding study procedures, risks, and confidentiality protections.

For hospitalised participants lacking decision-making capacity, consent will be sought from legally authorised representatives in accordance with Indonesian and UK ethical regulations. Participants who regain capacity during follow-up will be re-consented directly. Participants may withdraw from the study at any time without affecting their medical care.

Participants in studies involving direct participation will receive reimbursement for time and inconvenience in accordance with locally approved ethical guidance.

#### Data collection and management

Study data will be collected and managed using REDCap electronic data capture tools hosted at the Liverpool School of Tropical Medicine. REDCap (Research Electronic Data Capture) is a secure, web-based software platform designed to support data capture for research studies, providing 1) an intuitive interface for validated data capture; 2) audit trails for tracking data manipulation and export procedures; 3) automated export procedures for seamless data downloads to common statistical packages; and 4) procedures for data integration and interoperability with external sources (7, 8). For the INTERCEPT-Transmission study, demographic, clinical, antimicrobial exposure, hospitalisation, travel, and microbiological data will be collected from hospitalised participants, community participants, and healthcare workers. Weekly antimicrobial point prevalence surveys will also be conducted on participating wards to describe prescribing practices, including antimicrobial agents, doses, routes, indications, and planned duration. Participants will be assigned pseudonymised study identifiers, and identifiable information will be stored separately from research datasets with restricted access.

For the INTERCEPT-Bacteraemia study, anonymised clinical and microbiological data will be collected for patients with Gram-negative bloodstream infections, including demographics, comorbidities, antimicrobial treatment, hospitalisation details, and clinical outcomes.

For the INTERCEPT-Intervention study, qualitative data will be collected through semi-structured interviews, focus group discussions, and participant observation. Interviews and discussions will explore attitudes, perceptions, barriers, and practices related to antimicrobial stewardship and IPC implementation. The interview topic guides will be developed using the Theoretical Domains Framework (TDF) to ensure systematic exploration of behavioural determinants relevant to intervention development (9, 10). Ethnographic observations will be documented using reflexive field diaries to understand contextual and behavioural influences on stewardship and IPC practices, as well as intervention development processes (10). Qualitative interview recordings, transcripts, and ethnographic field notes will be anonymised prior to analysis. Genomic and microbiological datasets will be securely stored and managed according to institutional and national data governance requirements. Access to study data will be restricted to authorised study personnel.

#### Sample collection

In the INTERCEPT-Transmission study, stool or rectal swab samples will be collected longitudinally from hospital inpatients and once from community participants (Table 2). Hospital inpatients will be sampled at recruitment, then 3 and 7 days later and weekly while they remain an inpatient. Healthcare workers will undergo hand, clothing, and nasopharyngeal sampling twice during recruitment periods. Environmental samples, including high-touch surfaces, sinks, toilets, medical equipment, and wastewater, will be collected weekly from participating wards and hospital wastewater systems.

**Table 2.**
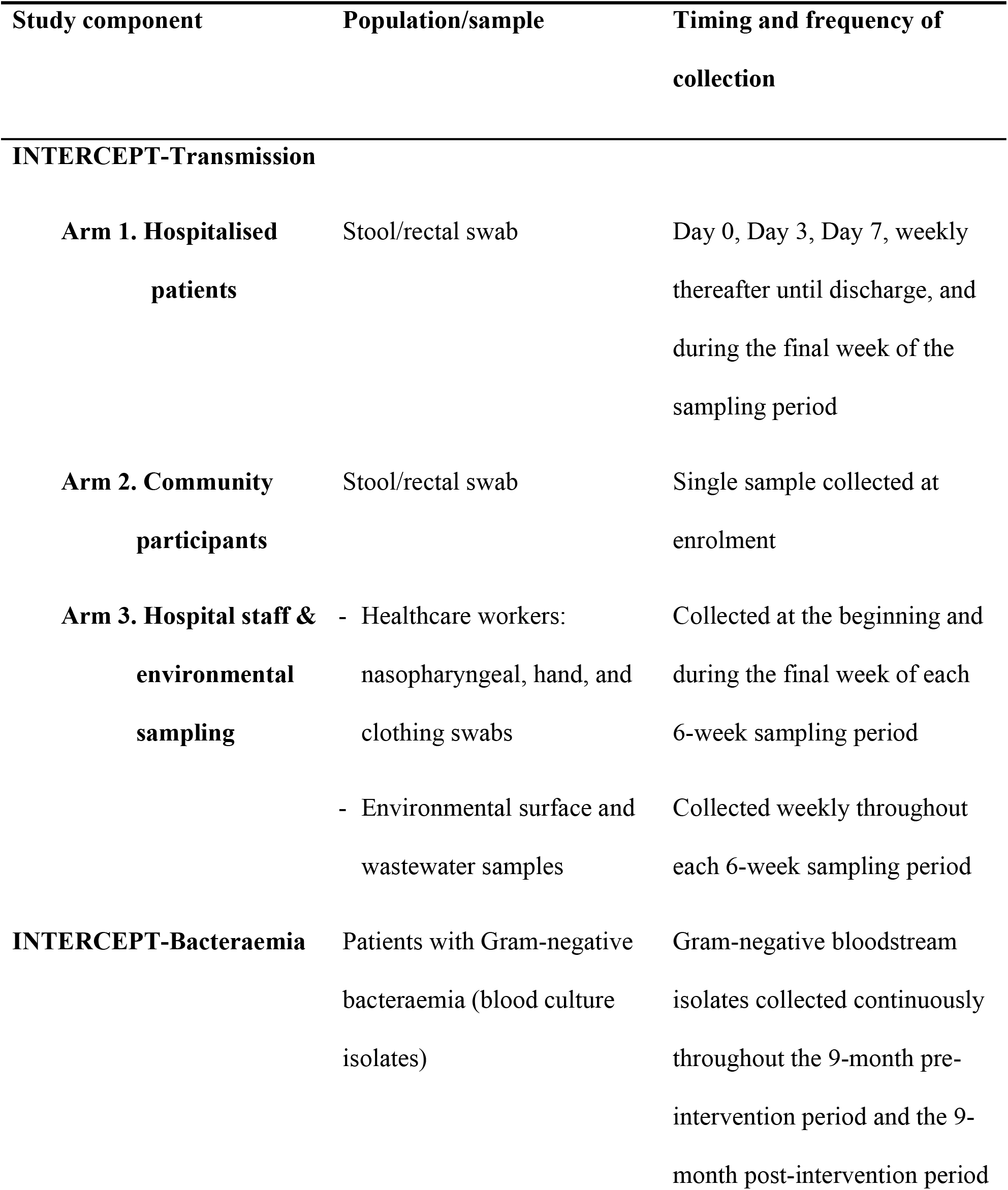
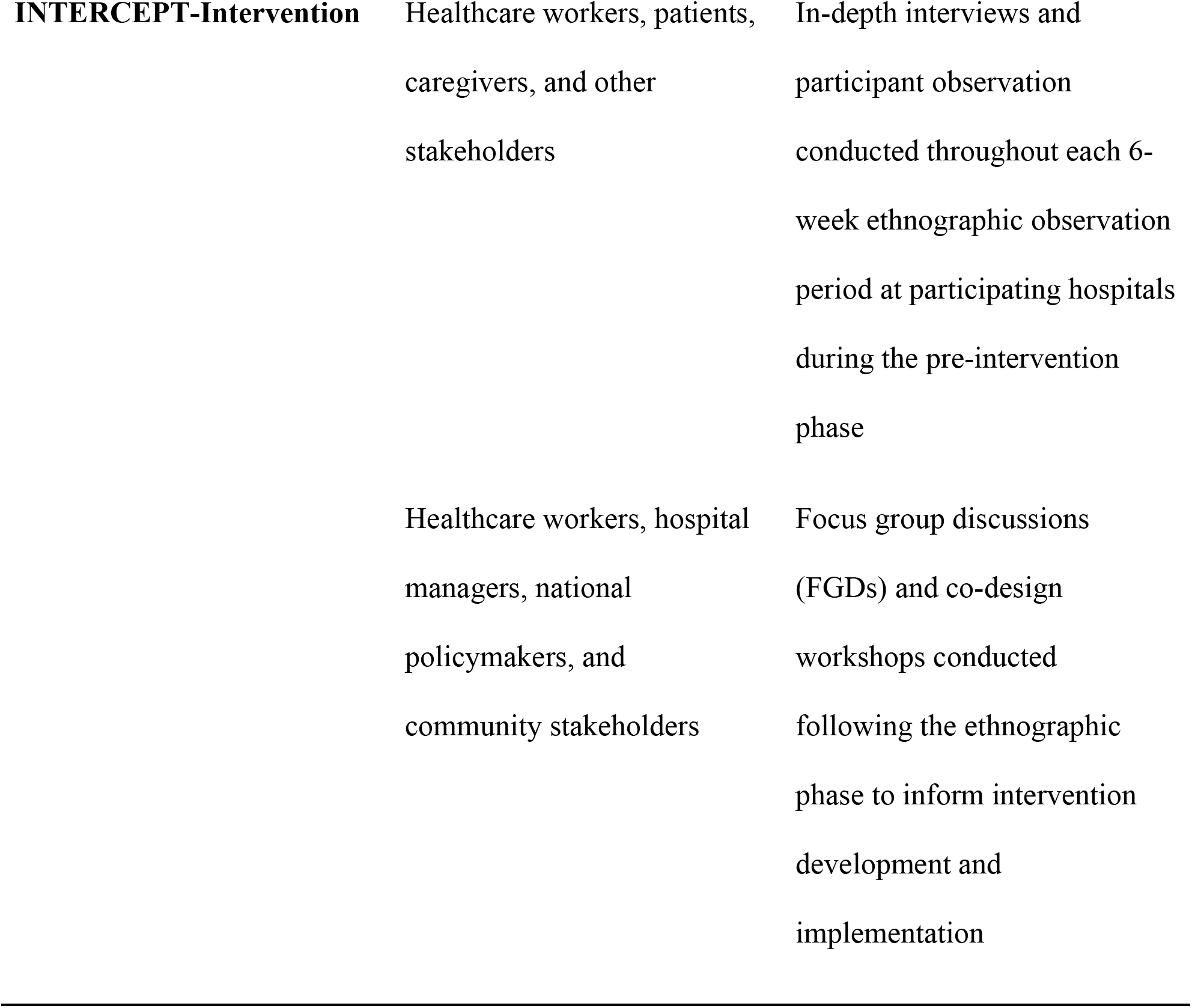
Sampling and data collection procedures across INTERCEPT study components.

#### Laboratory procedures

For the INTERCEPT-Transmission study, samples will undergo selective culture for carbapenem-resistant organisms followed by bacterial identification, antimicrobial susceptibility testing, and genomic analysis. Single isolate whole-genome sequencing using both short-and long-read platforms, alongside post-enrichment (‘plate sweep’) metagenomic sequencing (11, 12), will be performed to characterise bacterial strains, resistance genes, plasmids, and other mobile genetic elements. Selected isolates and extracted DNA may be transferred to collaborating laboratories outside Indonesia under approved material transfer agreements. To facilitate reproducibility while maintaining manuscript brevity, detailed laboratory protocols are provided separately. These protocols describe all microbiological, genomic, bioinformatic, and experimental procedures used in the INTERCEPT study. Detailed laboratory methods are provided in Supplementary Methods.

For the INTERCEPT-Bacteraemia study, bloodstream isolates identified through routine clinical microbiology laboratories will undergo genomic analysis using similar approaches.

No biological samples will be collected in the INTERCEPT-Intervention study.

#### Quantitative data analysis

For the INTERCEPT-Transmission study, mathematical transmission modelling will be used to investigate acquisition and transmission dynamics of carbapenem-resistant bacteria among hospitalised patients and healthcare environments. Stochastic multistate transmission models will estimate transitions between colonised and non-colonised states, with transition rates influenced by patient-level, environmental, antimicrobial exposure, and healthcare-associated factors (13, 14). Genomic and epidemiological data will be integrated into transmission analyses using pairwise genomic similarity approaches and within-host diversity estimates derived from limited-diversity metagenomic sequencing. These models will be used to identify likely transmission pathways and quantify the relative contribution of patients, healthcare workers, environmental surfaces, and wastewater systems to the spread of carbapenem-resistant bacteria.

Descriptive and regression-based analyses will be used to characterise colonisation prevalence, antimicrobial exposure, and transmission-associated risk factors. Community prevalence estimates will be calculated with corresponding confidence intervals.

Genomic and metagenomic data will be integrated with epidemiological and microbiological findings to investigate transmission pathways, resistance gene dissemination, plasmid dynamics, and mechanisms of horizontal gene transfer.

For the INTERCEPT-Bacteraemia study, descriptive analyses will summarise the microbiological and clinical characteristics of Gram-negative bloodstream infections. Regression analyses will compare outcomes between carbapenem-resistant and non-resistant infections, including mortality and length of hospital stay.

#### Qualitative data analysis

For the INTERCEPT-Intervention study, qualitative data from interviews, focus group discussions, ethnographic observations, field notes, and reflexive diaries will be analysed using the Framework Approach, supported by NVivo 12 software (QSR International, Melbourne, Australia). Data collection and analysis will be conducted iteratively, allowing emerging findings from early interviews and observations to inform subsequent data collection and refinement of interview guides.

An initial analytical framework will be developed based on the study objectives, the Theoretical Domains Framework (TDF), and recurring concepts identified during early familiarisation with the data. This framework will remain flexible to accommodate inductively emerging themes throughout the analysis (10).

Interview transcripts, focus group discussions, observational field notes, and reflexive diaries will be coded independently by members of the qualitative research team. Relevant codes will be grouped into categories and progressively refined into overarching themes. Once preliminary themes have been identified, transcripts and other qualitative data sources will be revisited using constant comparison to further develop, refine, or challenge emerging interpretations. Particular attention will be paid to atypical cases, conflicting accounts, and contradictory findings to retain the richness and complexity of participants’ experiences. Three researchers will independently code an initial subset of transcripts. Coding discrepancies will be discussed and resolved through consensus. Divergent cases, contradictory perspectives, and contextual factors will be actively explored to improve the credibility and richness of the analysis.

Findings from different qualitative data sources will be triangulated to strengthen interpretation and to provide a comprehensive understanding of behavioural, organisational, and contextual factors influencing antimicrobial stewardship and infection prevention and control practices. Regular discussions among the multidisciplinary research team will be undertaken throughout the analytical process to review emerging themes, resolve discrepancies in interpretation, and ensure analytical rigour.

The qualitative findings will subsequently inform stakeholder workshops for the co-design of context-specific antimicrobial stewardship and IPC interventions.

Missing quantitative data will be addressed using multiple imputation or Bayesian modelling approaches where appropriate.

#### Trustworthiness of qualitative findings

Credibility of the qualitative findings will be strengthened through triangulation of multiple qualitative data sources, iterative data collection and analysis, reflexive field notes, and regular discussions among the multidisciplinary research team to refine emerging interpretations. Sampling will continue until sufficient conceptual richness has been achieved for each stakeholder group.

#### Sample size calculation

For INTERCEPT-Transmission, we used a simulation-based approach to calculate the sample size, aiming to correctly identify factors that confer an increased colonisation pressure. Multistate models were constructed with an instantaneous rate of colonisation or decolonisation given by (0.01, 0.005) day-1 respectively, based on values from our previous study (13). Other colonised patients and the environment on the ward act to increase the rate of colonisation via a force of infection with a hazard ratio HR_1_ or HR_2,_ respectively, per 10% increase in proportion of colonised patients/ environmental sources. Initial ward patient colonisation proportion was drawn from a Uniform (0,0.1) distribution and environmental colonisation drawn from Uniform (0,0.8). Twelve wards were considered, varying the number of recruited participants (n) and the values of HR_1_/HR_2_, with 100 simulations for each combination of parameter values. Multistate models were fitted using the msm package in R, simulating csample collection for each patient at days 0, 3 and 7. The proportion of simulations in which the estimate of a given HR was correctly found to have 95% confidence intervals > 1 (i.e. a measure of the power of the study) are shown as heatmaps for environmental (Figure 2, left panel) and human-to-human (Figure 2, right panel) transmission; n > 500 is likely (∼ > 80% of simulations) identify HR equal to or greater than 1.3. Using this approach we estimate that 500 patients per sampling round (i.e. over 9 months, 1000 patients total) is likely to identify a clinically relevant effect on driving increase in colonisation (Figure 3). The primary outcome is the value of the parameter governing acquisition of carbapenem-resistant bacteria.

**Figure 2:**
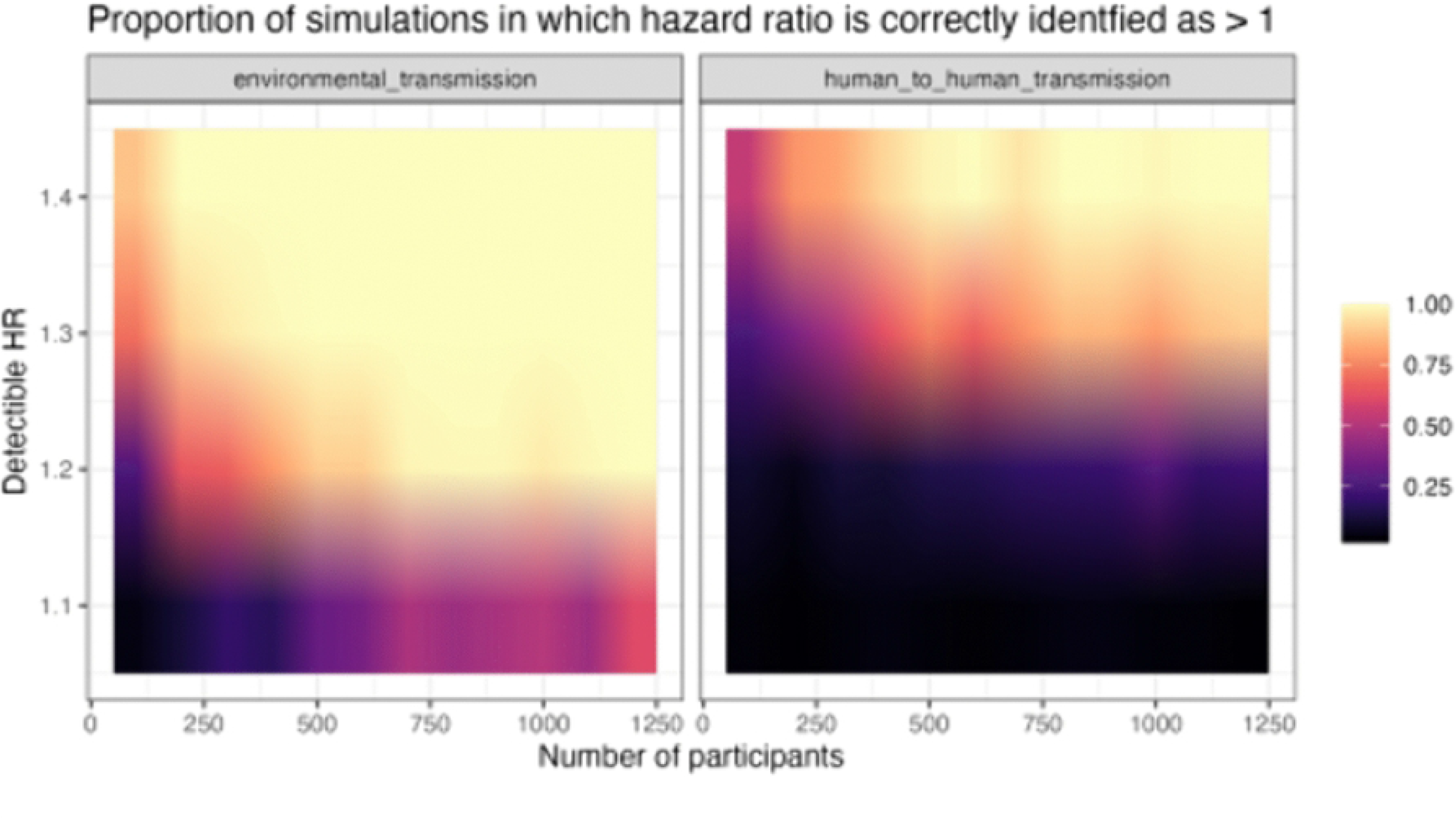
Sample size calculation using a simulation-based approach showing proportion of simulations in which a hazard ratio for increased colonisation pressure is correctly identified as number of participants (x axis) and detectable HR (y axis) varies.

Staff members will be incorporated in the models providing a force of infection term to the patients, where staff are colonised. No formal sample size calculation for staff has been carried out, but we aim to sample as many staff as possible for a given ward – we estimate that 20 staff per ward is feasible, resulting in 120 people in total.

100 community members recruited from the communities served by each hospital will (assuming community prevalence of colonisation 5%) result in 300 participants in total, and allow an estimation of the community prevalence of colonisation with carbapenem-resistant bacteria of +/-3%.

The INTERCEPT-Bacteraemia study will be a description of the clinical and bacteriologic features of Gram-negative bacteraemia. Anonymised patient metadata (age, gender, clinical outcome, length of stay) will be used to describe clinical features of bacteraemia. No sample size calculation is carried out for this descriptive cohort; 100 patients over 9 months (3 per week) is feasible.

For INTERCEPT-Intervention our general analytical approach will be developmental and iterative, in that data collection and analysis will be undertaken concurrently; knowledge generated in the early interviews and focus groups will be reviewed and compared with and built on in subsequent interviews and observational ethnography. This approach will provide an opportunity to establish initial findings and build on these as data collection evolves. We will develop summary tables of emerging findings and continuously discuss these within the wider research team. This will enable initial findings to be challenged, revealing relevant gaps that will make subsequent data collection efficient.

### Ethics

Ethical approvals have been obtained from the Liverpool School of Tropical Medicine Research Ethics Committee (approval number: 25-041) and the Faculty of Medicine, Universitas Diponegoro Ethics Committee (approval number: 312/EC/KEPK/FK-UNDIP/X/2025). Additional ethical approvals have been obtained from the participating hospitals: Dr. Kariadi Central Hospital Ethics Committee (approval number: 16519/KEPK-RSDK/2025) and K.R.M.T. Wongsonegoro District Hospital Ethics Committee (approval number: 167/Kom.EtikRSWN/ VIII/2025). Dr. M. Ashari Hospital does not have an institutional research ethics committee and accepted the ethical approval granted by the Dr. Kariadi Central Hospital Ethics Committee for the conduct of this study at its site.

Written informed consent will be obtained from all participants who are directly recruited into the study or from their legally authorised representatives. The INTERCEPT-Bacteraemia component does not involve direct participant recruitment or additional patient procedures and will use anonymised routinely collected clinical and microbiological data and bacterial isolates obtained through routine clinical microbiology services. Individual informed consent will not be obtained for this component.

Participant confidentiality will be protected through pseudonymisation and secure data storage. Biological materials transferred outside Indonesia for genomic or molecular analysis will be managed under approved material transfer agreements and relevant national regulations.

## Discussion

Carbapenem-resistant bacteria represent one of the most urgent antimicrobial resistance threats globally, particularly in low-and middle-income countries where healthcare systems face major diagnostic, therapeutic, and infection prevention challenges. Indonesia has experienced increasing rates of carbapenem resistance despite implementation of national antimicrobial resistance control programmes and antimicrobial stewardship initiatives (3, 15). However, important knowledge gaps remain regarding the transmission dynamics of carbapenem-resistant organisms across Indonesian healthcare and community settings. The INTERCEPT study aims to address these gaps through an integrated multidisciplinary approach combining microbiological surveillance, genomics, mathematical modelling, qualitative research, and implementation-focused intervention development.

A major strength of the INTERCEPT study is its systems-level approach to understanding antimicrobial resistance transmission. Rather than focusing solely on clinical infection, the study investigates interconnected pathways linking hospitalised patients, healthcare workers, hospital environments, wastewater systems, and surrounding communities. This approach recognises that carbapenem resistance is driven not only by antimicrobial selection pressure, but also by transmission across multiple ecological and healthcare domains. By incorporating environmental and community sampling alongside clinical surveillance, the study seeks to provide a more comprehensive understanding of how resistant organisms and resistance genes circulate within and beyond healthcare facilities.

Another important strength is the integration of whole-genome sequencing and mathematical transmission modelling. Previous studies in Indonesia have largely relied on phenotypic resistance data or cross-sectional microbiological surveillance. In contrast, INTERCEPT combines short-and long-read genomic sequencing with transmission modelling to investigate both bacterial dissemination and movement of mobile genetic elements, including plasmids carrying carbapenemase genes. This integrated analytical approach may help identify dominant transmission pathways and provide evidence to guide targeted infection prevention and antimicrobial stewardship interventions.

The study also incorporates qualitative and ethnographic methods to better understand behavioural and system-level barriers influencing IPC and antimicrobial stewardship implementation. Although IPC and antimicrobial stewardship programmes have been introduced in many Indonesian hospitals, evidence regarding implementation challenges, contextual barriers, and sustainability remains limited. Through interviews, focus group discussions, participant observation, and stakeholder engagement workshops, INTERCEPT seeks to ensure that proposed interventions are contextually appropriate and feasible within diverse Indonesian healthcare settings.

Importantly, the intervention component of the study adopts a co-design approach involving healthcare workers, hospital managers, policymakers, and community stakeholders. This participatory strategy may improve local ownership, acceptability, and long-term sustainability of interventions. The inclusion of tertiary, secondary, and district-level hospitals also enables assessment of transmission dynamics and intervention feasibility across different levels of the Indonesian healthcare system.

This study has several limitations. First, recruitment is limited to selected wards and hospitals in Central Java and therefore may not fully represent transmission dynamics across all Indonesian healthcare settings. Second, although genomic and modelling approaches may strengthen inference regarding transmission pathways, unsampled sources and incomplete sampling are likely. Third, intervention evaluation will primarily focus on feasibility and implementation outcomes rather than definitive effectiveness outcomes. These will need to be assessed in larger cluster-randomised studies.

Despite these limitations, INTERCEPT is expected to provide one of the most comprehensive investigations of carbapenem resistance transmission conducted in Indonesia to date. Findings from this study may inform national antimicrobial resistance control strategies, support development of locally adapted IPC and antimicrobial stewardship interventions, and strengthen research capacity in genomics, bioinformatics, and implementation science. The integrated methodological framework developed through INTERCEPT may also be applicable to other low-and middle-income settings facing similar antimicrobial resistance challenges.

## Data Availability

No datasets were generated or analysed for this study protocol. All relevant data from our study will be made available upon study completion, in accordance with applicable ethical, legal, and institutional requirements.

## Supplementary materials

S1 File. Stool Rectal Swab Sampling

S2 File. Stool Sample Processing

S3 File. Environmental Swab Sampling

S4 File. Environmental Swab Processing

## Acknowledgements

The authors acknowledge the support provided by the Ministry of Education, Culture, Research and Technology of the Republic of Indonesia (Kemendikbudristek) and the Indonesia Endowment Fund for Education (LPDP) for the Indonesian component of the study.

The authors would also like to thank the participating hospitals, healthcare workers, laboratory staff, community participants, and policymakers involved in the development of the INTERCEPT study. We also acknowledge the contributions of the wider INTERCEPT collaborative research team in Indonesia and the United Kingdom.

## AI Disclosure

Generative AI tools were used to assist language refinement and manuscript editing under author supervision. All scientific content was reviewed and approved by the authors.

## Author contributions

Conceptualisation: HF, RH, ESL, NF, AR, FEG, RD, MM, JML

Methodology: HF, RH, ESL, NF, AR, FEG, RD, MM, JML

Investigation: HF, RH, ESL, NF, AR, FEG, RD, MM, JML

Formal analysis: JML

Supervision: HF, RH, ESL, NF, AR, FEG, RD, MM, JML

Writing – original draft: HF

Writing – review & editing: HF, RH, ESL, NF, AR, FEG, RD, MM, JML

## Data availability statement

This Study Protocol does not report data; therefore, the PLOS Data Availability Policy is not currently applicable. Study findings will be disseminated through peer-reviewed publications, conference presentations, stakeholder workshops, and engagement with healthcare institutions and policymakers. Results will also be shared with participating hospitals and communities where appropriate.

Following publication, anonymised datasets, analytical code, and genomic sequence data will be made available through appropriate repositories, subject to ethical and regulatory requirements, in accordance with the Findable, Accessible, Interoperable, and Reusable (FAIR) principles (16).

